# Phenome-wide association study to explore the long-term symptoms after infection with novel coronavirus in the UK Biobank

**DOI:** 10.1101/2023.03.03.23286122

**Authors:** Kai Zhang, Xiaowen Liu, Ping Fu, Yingqi Zhao, Qingqing Yu, Shuling Liu, Fuzhong Xue

**Author notes:** **Correspondence to:** Fuzhong Xue, Ph.D., Department of Biostatistics, School of Public Health, Cheeloo College of Medicine, Shandong University, No.44 Wenhuaxi Road, Jinan, 250012, China.).

## Abstract

**Background:** Observational research studies have shown that even after the acute phase, severe acute respiratory syndrome coronavirus 2 (SARS-CoV-2) can affect patients, and increase the risk of cardiovascular, mental, metabolic, and other disorders. However, the spectrum of diseases for individuals with a genetic predisposition to COVID-19 remains unclear.

**Methods:** We leveraged individual-level data from UK Biobank to implement a phenome-wide association study to explore the relationships between COVID-19 and 1061 diseases. Then, the inverse-variance weighted (IVW) method was adopted with summary-level data from global consortiums as sensitivity analyses combined with other MR methods with different model assumptions to identify robust associations.

**Findings:** The PheWAS found severe respiratory, hospitalized, and susceptibility COVID-19 had detrimental effects on 36, 37, and 51 kinds of diseases, separately. The IVW test found severe respiratory COVID-19 had detrimental effects on breast cancer [OR 95% CI: 1.065 (1.000-1.133)], pan-cancer [OR 95% CI: 1.002 (1.000-1.004)], and Alzheimer’s disease [OR 95% CI: 1.042 (1.005-1.081)], etc. Hospitalized COVID-19 had detrimental effects on ischemic stroke (IS) [OR 95%CI: 1.049 (1.001-1.100)], breast cancer [OR 95%CI: 1.139 (1.011-1.283)], and pan-cancer [OR 95%CI: 1.003 (1.000-1.006)], etc. Susceptibility COVID-19 had detrimental effects on deep vein thrombosis (DVT) of lower extremities [OR 95%CI: 2.392 (1.167-4.902)], venous thromboembolism [OR 95%CI: 1.962 (1.115-3.453)], pulmonary heart disease/diseases of pulmonary circulation [OR 95%CI: 1.767 (1.142-2.733)], IS (large artery atherosclerosis) [OR 95%CI: 1.405 (1.025-1.927)], myocardial infarction [OR 95%CI: 1.235 (1.012-1.509)], heart failure [OR 95%CI: 1.140 (1.009-1.287)], etc.

**Interpretation:** This study describes the extensive link between genetically determined COVID-19 and a broad range of diseases, especially those of the circulatory system, neuropsychiatric system, neoplasms, immune system, and digestive systems. Early detection and management of post-COVID-19 conditions could be tremendously beneficial to public health.

**Funding:** This work was supported by the National Natural Science Foundation of China (81773547 and 82173625) and the National Key Research and Development Program (2020YFC2003500).

## 1. Introduction

The coronavirus disease 2019 (COVID-19) has spread across the world. As of 31 December 2022, more than 657 million confirmed cases of COVID-19 have been recorded worldwide, and more than 6.68 million deaths have been reported by the World Health Organization (WHO). Much research shows that even after the acute phase, severe acute respiratory syndrome coronavirus 2 (SARS-CoV-2) can affect patients and impair their quality of life. Recently, International organizations and Institutes have drawn attention to an increasing number of people experiencing health consequences following the acute phase of SARS-CoV-2 infection and are calling for research into the risk factors, clinical features, diagnosis, management, and outcomes [1–5]. Indeed, even the terminology of the condition is debated with variable terms and definitions for the post-COVID-19 condition including long COVID, long-haul COVID, long COVID-19 condition, post-acute sequelae of SARS-CoV-2 infection (PASC), or post-COVID-19 condition (the term used by WHO) [6–8].

Long-term health consequences of COVID-19 remain unknown, but most study report that the spectrum of long-lasting symptoms is wide and varies from mild discomfort to severe adverse effects on physical, cognitive, and psychosocial health, with important wider implications on functioning, including employment and school attendance [9]. Multiple studies from different countries found that many individuals experienced persistent symptoms 6 months after COVID-19, with fatigue or muscle weakness, sleep difficulties, and anxiety or depression among the most common sequelae [10–12]. A recent study suggests that although most COVID-19 survivors recover both physically and functionally a year after acute infection, some still experience problems with mobility, pain or discomfort, and anxiety or depression compared with non-COVID-19 controls [13]. The data emerging from the controlled studies are in agreement with the earlier reports. A recent analysis of the data from over 87 million electronic health records demonstrated that more than one in three individuals had one or more features of post-COVID-19 condition recorded between 3 and 6 months after a diagnosis of COVID-19, which was significantly higher when compared with individuals with influenza [14–18]. Disease severity, female sex, and age were associated with a higher risk of post-COVID-19 condition development.

However, observational evidence is less likely to account for unknown confounding factors and reverse causation bias [19]. It is also important to note that most data regarding post-COVID-19 conditions have been generated before the condition definition announcement. Thus, earlier studies may not fit the proposed definition criteria. In addition, these studies have often focused on a single type of disease. The wide range of diseases associated with COVID-19 requires an urgent systematic assessment.

A phenome-wide association study (PheWAS) [20] combined with a two-sample Mendelian randomization (MR) approach was suitable for use in examining genetic evidence between the COVID-19 phenotypes and diseases, providing a more comprehensive assessment of post-COVID-19 condition associated human complex diseases. The PheWAS paradigm was introduced in 2010 as an approach that scans across a range of phenotypes, similar to what was accomplished in genome-wide association studies (GWAS) [20]. Studies that adopted PheWAS analysis have been used to understand the beneficial or detrimental effects of body mass index (BMI), serum urate, age at menarche, smoking intensity, ABO blood groups, and C-reactive protein (CRP) on human complex phenotypes [21–26].

This study leveraged both PheWAS approaches and MR to identify the spectrum of human diseases associated with genetically predisposed COVID-19. In addition, several complementary two-sample MR approaches were used to identify robust associations.

## 2. Materials and methods

### 2.1 Study Population

UK Biobank contains in-depth genetic and health information on over 500,000 participants.[27] This prospective study collected data including blood, urine, and saliva laboratory tests and questionnaires, physical measurements, and genome-wide genotyping information.[27] This information is linked to national primary care and inpatient records, cancer diagnosis, and death registration data for longitudinal follow-up.[27] Participants who withdrew from the study or were lost to follow-up, those lacking genotype information, self-reported sex data that did not match genetic records, sex chromosome aneuploidy, >10 putative third-degree relatives in the kinship table, excessive heterozygosity (top 1%), non-Caucasian ancestry, and missing key covariates (age, sex, BMI, smoke, and drink status) were excluded from the analysis. Genetic quality control was performed centrally by the UK Biobank.[28] Detailed information about this cohort can be found on the Biobank website (https://www.ukbiobank.ac.uk/). For PheWAS, 455,509 participants with genotypes in the UK Biobank were included.

### 2.2 GWAS summary-level data for COVID-19

We evaluated three COVID-19 phenotypes: severe respiratory, hospitalized, and susceptibility to COVID-19. These data were derived from Release 7 of the GWAS meta-analysis conducted by the COVID-19 Host Genetics Initiative (HGI). (https://www.covid19hg.org/results/r7/) This is an international genetics collaboration that aims to uncover the genetic determinants of COVID-19 susceptibility, severity, and outcomes.[29] For the evaluated COVID-19 outcomes, controls were genetically ancestry-matched individuals without SARS-CoV-2 infection.[29] In our analyses, we used GWAS summary statistics from the comparison between cases and control groups of each exposure. The severe respiratory COVID-19 outcome resulted from the comparison between patients with very severe respiratory failure secondary to COVID-19 (n = 18,152) vs controls (n = 1,145,546). The hospitalized COVID-19 data were generated from the comparison of patients with a laboratory-confirmed SARS-CoV-2 infection that was hospitalized due to COVID-19 symptoms (n = 44,986) vs controls (n = 2,356,386). Finally, the susceptibility COVID-19 analysis was conducted by comparing 159,840 individuals reporting SARS-CoV-2 infection with 2,782,977 controls. Information regarding SARS-CoV-2 infection was derived from a laboratory test, electronic health record, clinically confirmed COVID-19, and self-reported COVID-19 (e.g. by questionnaire).[29] Single nucleotide polymorphisms (SNPs) were filtered according to the following criteria: 1) a genome-wide threshold of significance (*P*<5×10^−8^) and 2) linkage disequilibrium (LD) defined as *r*^*2*^ < 0.01 and clump window > 10,000 kb.[30] **(Supplementary Table S1**)

### 2.3 Phenome Construction

Phenotypes were constructed using the PheCODE system, which was developed to combine one or more relevant International Classification of Diseases (ICD) codes into different disease groups, allowing unbiased interrogation of multiple phenotypes in an EMR-based cohort.[31] To construct the phenotypes, a map designed for large biobanks was used to match ICD-9/10 codes to the “phecode.”[32] The PheCODE system also provided a scheme to automatically exclude patients with similar or related diseases from the controls.[22] Primary and secondary ICD-9/10 codes in the UK Biobank were pooled from hospital records, cancer registry, and death registry data, and mapped into phecodes using the R package, “PheWAS”.[33] The phecode-mapping rules, as well as the excluding standards for each code, are available at https://phewascatalog.org/phecodes_icd10. Phecodes with more than 200 cases were included in the final analysis. A total of 1061 phenotypes were utilized in the PheWAS and divided into 17 categories: circulatory, congenital, dermatologic, digestive, endocrine/metabolic, genitourinary, hematopoietic, infectious disease, injuries and poisonings, mental disorders, musculoskeletal, neoplasms, neurological, pregnancy complications, respiratory, sense organ, and symptom disorders. **(Supplementary Table S2**)

### 2.4 Statistical Analyses

#### 2.4.1 Phenome-wide association study

The PRS was determined using the equation below.

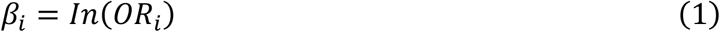

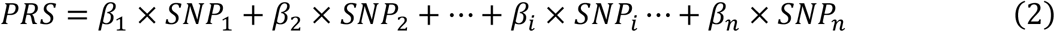

This PRS calculation method assumes an additive genotype model that an individual may have 0, 1, or 2 risk-increasing alleles. The sum of the risk alleles is calculated using the effect size from the variant-exposure association, defined as *β*, which is obtained from the discovery of GWAS [34], and *n* is defined as the total number of SNP included in the model. *Z*-transformation was performed on GRS to ensure that the final result corresponded to the disease risk of 1-SD PRS increments. All models assumed a multivariable logistic model and adjusted for age, sex, BMI, smoking status, and drinking status. The “PheWAS” package was applied to test associations between genetically determined COVID-19 and hundreds of phenotypes in the UK Biobank cohort. The signals that passed the Bonferroni correction in the PheWAS analysis were used for further two-sample MR analyses.

#### 2.4.2 Two-Sample Mendelian Randomization

For the candidate phenotypes detected using PheWAS, two-sample MR analyses were performed in parallel to assess the robustness of causal findings. Summary-level datasets for these diseases were acquired from global consortiums and excluded the UK Biobank-related data sources to avoid potential bias from sample overlap. A total of 207 disease phenotypes in the finding phase have available GWAS datasets. These summary-level statistics was shown in **Supplementary Table S3**. The *F*-statistic of >10 could be considered a strong IV.

The IVW approach was used as the primary analysis with the assumption of no invalid IVs.[35] When the number of IVs was less than or equal to three, we applied the fixed-effects model to estimate the causal effect between exposure and outcome, otherwise, the random-effects model was used. Furthermore, the simple median (SME) and weighted-median estimator (WME) allow more powerful genetic variants to contribute more. It could be obtained by weighting the contribution of each genetic variant according to the inverse variance associated with the result. Even if up to 50% of the IVs are invalid, the estimator is consistent. [36] MR-Egger evaluates whether the pleiotropic effect of genetic variants on the result is different from zero on average [37]. MR-Egger is similar to IVW, but the former adjusts IVW analysis by allowing non-zero intercepts, namely, allowing horizontal pleiotropic effects. Even if all of the genetic variants violate IV assumptions 2, MR-Egger also returns an unbiased estimate of causal effects. [37] In MR-Egger regression, the estimate of intercept can be interpreted as an estimate of the average pleiotropy of all genetic variants, and the slope coefficient provides an estimate of the bias of the causal effect. [37] For binary variables, the MR estimates were reported as odds ratios (ORs) which can be interpreted as the risk increase of outcome per unit increase in log odds of exposure.

Phenotypes that passed the sensitivity analysis were defined as robust associations. All analyses were two-tailed and performed using R software (Version 3.6.3) with the ‘phewas’ and ‘TwoSampleMR’ packages.

## 3. Results

**Figure 1A and Figure 1B** showed the results of the PheWAS analysis between severe respiratory COVID-19 and 1061 binary phenotypes in the UK Biobank. **Figure 1A** showed that severe respiratory COVID-19 had detrimental effects on 36 diseases. These diseases were categorized as neoplasms (n=6), congenital anomalies (n=5), circulatory system (n=3), dermatologic (n=3), digestive (n=3), infectious diseases (n=3), endocrine/metabolic (n=2), injuries & poisonings (n=2), musculoskeletal (n=2), and one kind of mental disorders, neurological, respiratory, sense organs, symptoms, respectively. No associations were evident between severe respiratory COVID-19 and hematopoietic disorders and pregnancy complications.

**Figure 1.**
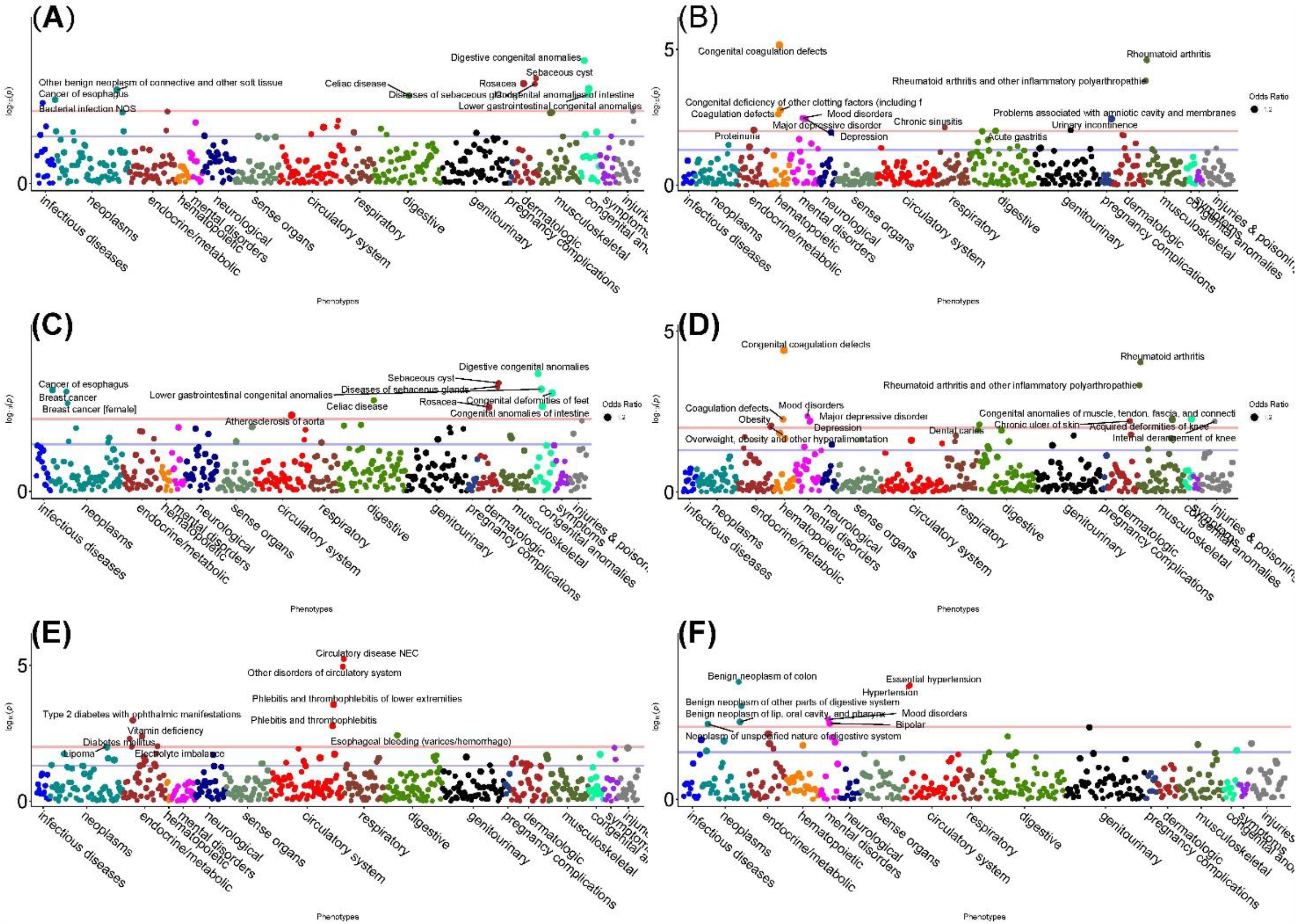
The results of PheWAS analysis in UK Biobank. The blue line indicates the false discovery rate (FDR) threshold. The red line indicates the self-defining notable threshold (*P*_*PheWAS*_ <0.01) to facilitate ideal visualization. Y-axis is minus log transformed *P*-value of the association between COVID-19 polygenic risk score (PRS) and disease outcomes; the X-axis provides the list of labels of 17 diseases category.

The odds ratios are shown in **Figure 2**. For circulatory system diseases, the odds ratio for the association between severe respiratory COVID-19 and atherosclerosis of aorta, other specified cardiac dysrhythmias, varicose veins of lower extremity, varicose veins were 1.160 (95% Confidence interval, 95% CI,1.028−1.293), 1.033 (95% CI,1.001−1.065), 1.022 (95% CI,1.004−1.040), and 1.020 (95% CI,1.002−1.037), respectively. For congenital anomalies, the odds ratios for the association between severe respiratory COVID-19 and congenital anomalies of intestine, lower gastrointestinal congenital anomalies, congenital deformities of feet, digestive congenital anomalies, upper gastrointestinal congenital anomalies were 1.199 (95% CI, 1.082−1.317), 1.151 (95% CI, 1.057−1.245), 1.137 (95% CI, 1.016−1.259), 1.127 (95% CI, 1.061−1.194), and 1.101 (95% CI, 1.007−1.195), respectively. For neoplasms, the odds ratios for the association between severe respiratory COVID-19 and other benign neoplasm of connective and other soft tissue, cancer of esophagus, benign neoplasm of brain and other parts of nervous system, cancer of stomach, benign neoplasm of brain/cranial nerves/meninges, breast cancer, encephalitis were 1.124 (95% CI, 1.048−1.199), 1.092 (95% CI, 1.031−1.153), 1.078 (95% CI, 1.020−1.135), 1.075 (95% CI, 1.007−1.144), 1.066 (95% CI, 1.006−1.126), 1.021 (95% CI, 1.001−1.041), and 1.104 (95% CI, 1.006−1.202), respectively.

**Figure 2.**
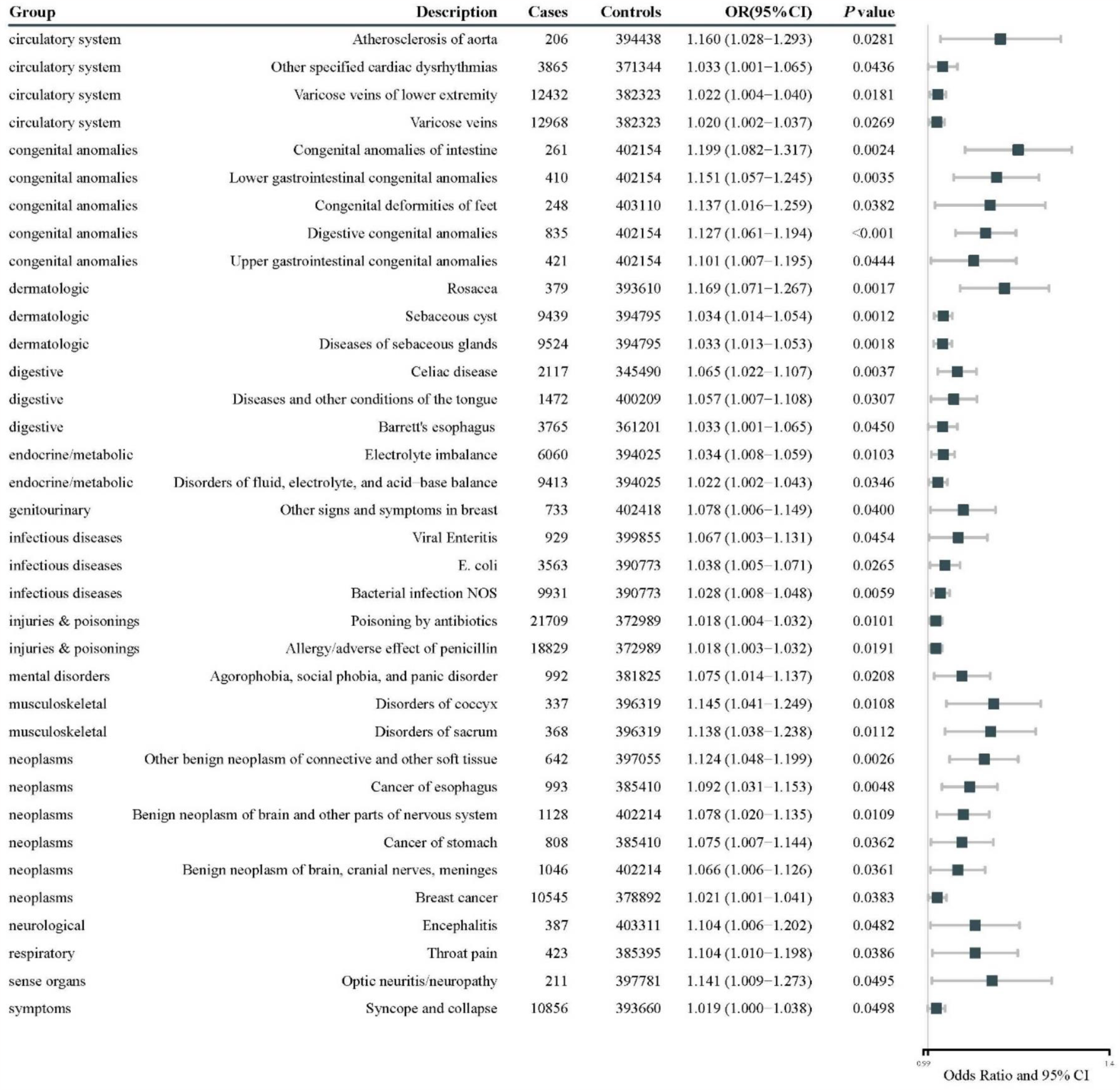
The odds ratios of PheWAS analysis between severe respiratory COVID-19 and diseases. Some diseases noted with ‘other’ and ‘un-specified’ was not shown in this Figure but displayed in the supplement material; OR: odds ratio; CI: Confidence interval.

**Figure 1C** showed that hospitalized COVID-19 had detrimental effects on 37 diseases. These diseases were categorized as neoplasms (n=7), congenital anomalies (n=4), genitourinary(n=4), circulatory system (n=3), dermatologic (n=3), digestive (n=3), endocrine/metabolic (n=3), injuries & poisonings (n=2), musculoskeletal (n=2), neurological (n=2), mental disorders (n=1), respiratory (n=1), and sense organs (n=1). The odds ratios are shown in **Figure 3**. For circulatory system diseases, the odds ratio for the association between hospitalized COVID-19 and cancer of esophagus, other benign neoplasm of connective and other soft tissue, breast cancer, breast cancer [female], lipoma of skin and subcutaneous tissue, cancer of prostate, malignant neoplasm of female breast were 1.104 (95% CI, 1.043−1.165), 1.087 (95% CI, 1.010−1.163), 1.032 (95% CI, 1.012−1.051), 1.030 (95% CI, 1.010−1.049), 1.029 (95% CI, 1.001−1.057), 1.025 (95% CI, 1.005−1.046), and 1.023 (95% CI, 1.001−1.044), respectively. For circulatory system, the odds ratio for the association between hospitalized COVID-19 and atherosclerosis of aorta, varicose veins of lower extremity, varicose veins were 1.197 (95% CI, 1.064−1.329), 1.021 (95% CI, 1.004−1.039), and 1.019 (95% CI, 1.001−1.036), respectively.

**Figure 3.**
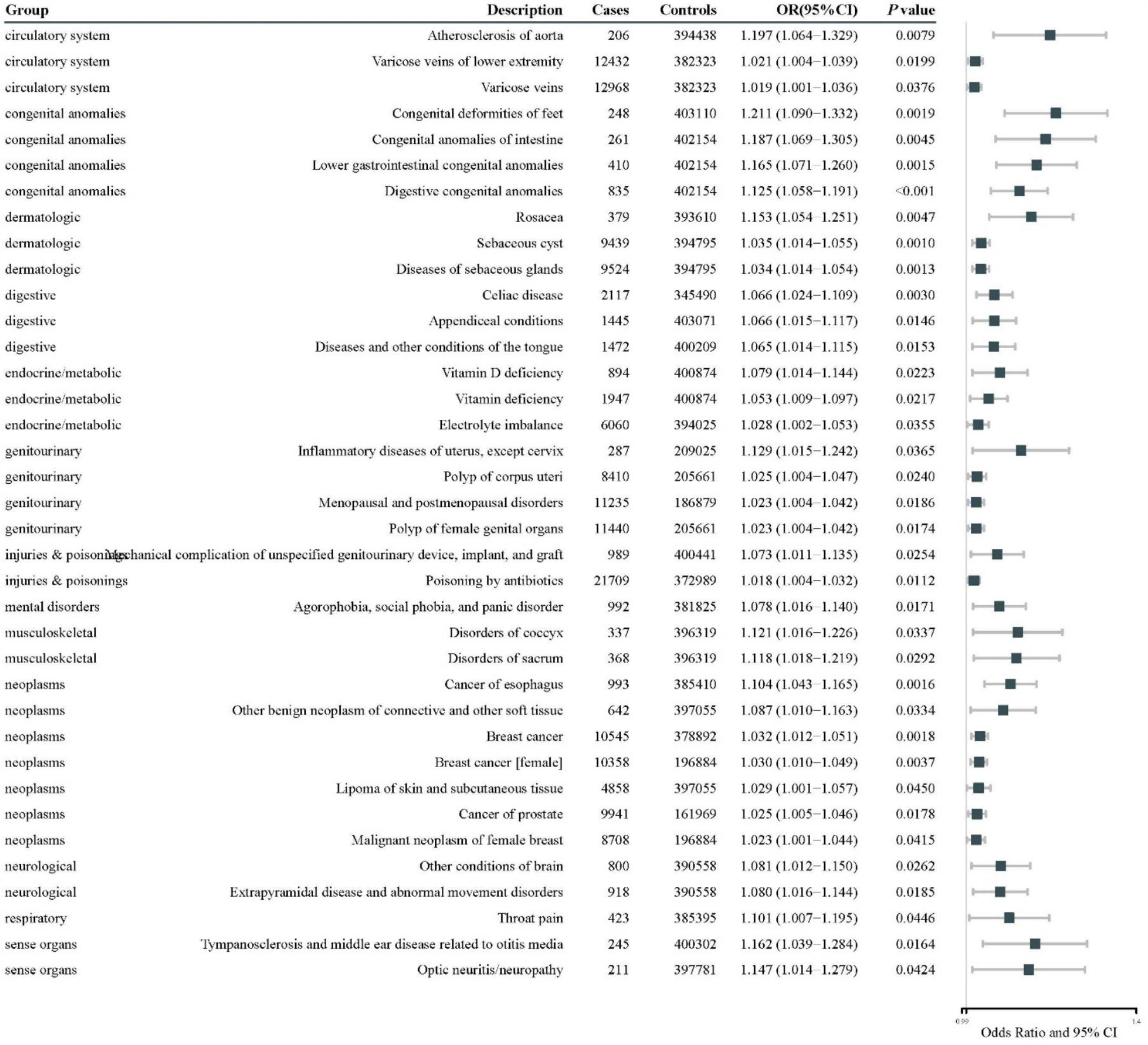
The odds ratios of PheWAS analysis between hospitalized COVID-19 and diseases. Some diseases noted with ‘other’ and ‘un-specified’ was not shown in this Figure but displayed in the supplement material; OR: odds ratio; CI: Confidence interval.

**Figure 2E** showed that susceptibility to COVID-19 had detrimental effects on 51 diseases. These diseases were categorized as endocrine/metabolic (n=10), circulatory system (n=9), neoplasms (n=7), dermatologic (n=6), digestive (n=4), musculoskeletal (n=3), respiratory (n=3), genitourinary (n=2), sense organs (n=2), symptoms (n=2), injuries & poisonings (n=1), neurological (n=1), and congenital anomalies (n=1). The odds ratios are shown in **Figure 4**. For endocrine/metabolic system diseases, the odds ratio for the association between COVID-19 infection and mineral deficiency, crystal arthropathies, type 2 diabetes with ophthalmic manifestations, vitamin D deficiency, vitamin deficiency, vitamin B−complex deficiencies, electrolyte imbalance, disorders of fluid/electrolyte/ and acid−base balance, diabetes mellitus, type 2 diabetes were 1.118 (95% CI, 1.016−1.221), 1.097 (95% CI,1.007−1.186), 1.096 (95% CI, 1.041−1.151), 1.078 (95% CI, 1.013−1.143), 1.067 (95% CI, 1.023−1.111), 1.063 (95% CI, 1.003−1.122), 1.034 (95% CI, 1.009−1.060), 1.025 (95% CI, 1.005−1.046), 1.020 (95% CI, 1.006−1.034), and 1.019 (95% CI, 1.005−1.033), respectively. For circulatory system, the odds ratio for the association between COVID-19 infection and atherosclerosis of aorta, other venous embolism and thrombosis, phlebitis and thrombophlebitis of lower extremities, phlebitis and thrombophlebitis, cardiac arrest and ventricular fibrillation, circulatory disease, other disorders of circulatory system, precordial pain, cardiac dysrhythmias, other specified congenital anomalies of kidney were 1.164 (95% CI, 1.031−1.296), 1.153 (95% CI, 1.034−1.271), 1.117 (95% CI, 1.057−1.177), 1.085 (95% CI, 1.034−1.136), 1.068 (95% CI, 1.007−1.129), 1.034 (95% CI, 1.019−1.048), 1.032 (95% CI, 1.018−1.046), 1.032 (95% CI, 1.002−1.061), 1.016 (95% CI, 1.004−1.028), and 1.121 (95% CI, 1.025−1.216), respectively.

**Figure 4.**
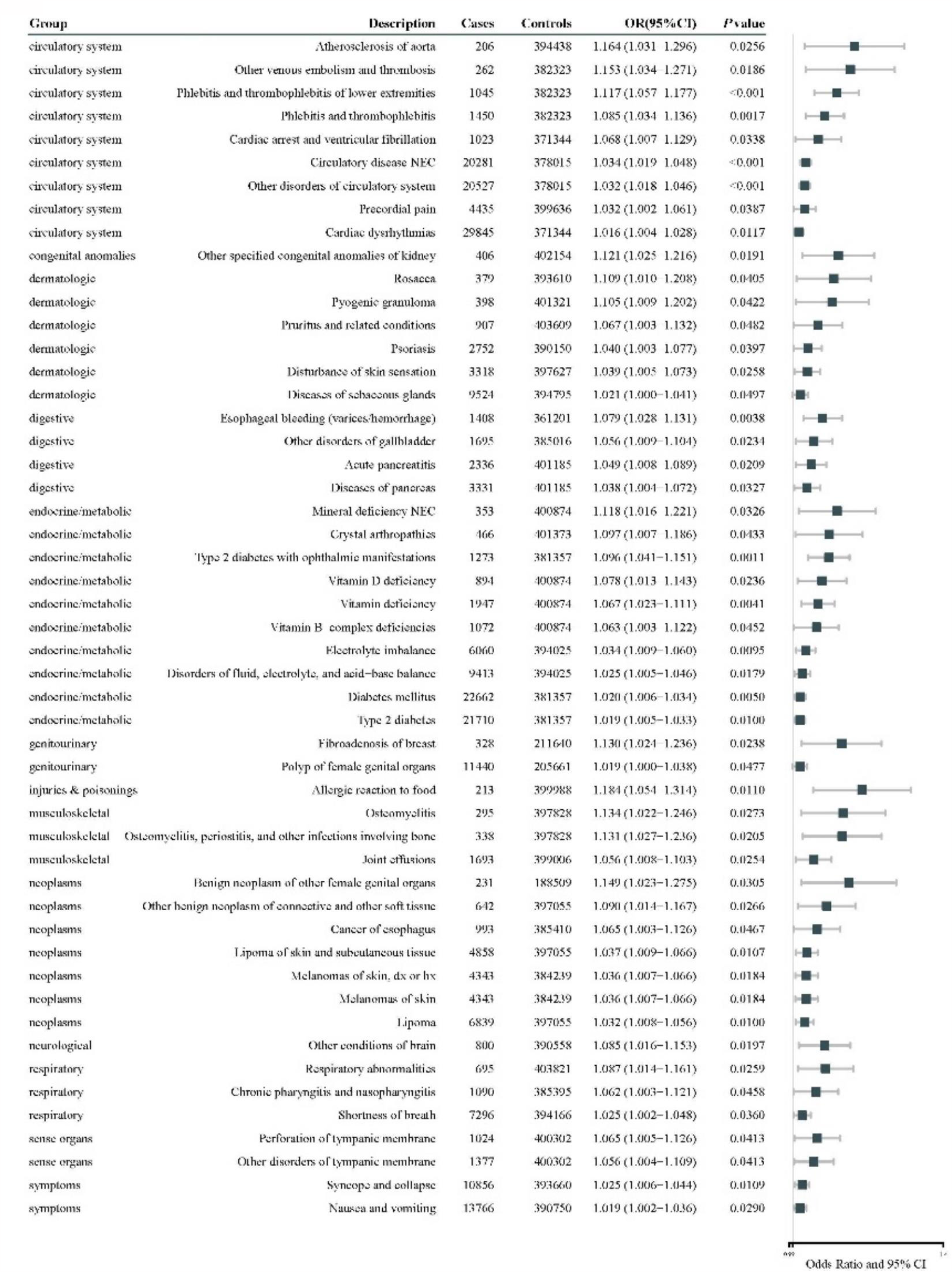
The odds ratios of PheWAS analysis between susceptibility COVID-19 and diseases. Some diseases noted with ‘other’ and ‘un-specified’ was not shown in this Figure but displayed in the supplement material; OR: odds ratio; CI: Confidence interval.

As shown in **Figure 5**, IVW test found a statistically significant association between critical COVID-19 and 26 diseases, including circulatory system (n=3): ischemic stroke (small−vessel) (OR, 95% CI: 0.941, 0.887-0.998), conduction disorders (OR, 95% CI: 0.935, 0.880 -0.992), atrioventricular (AV)-block (OR, 95% CI: 0.900, 0.830-0.976); digestive system (n=5): gastroesophageal reflux disease (OR, 95% CI: 1.022, 1.004-1.040), hernia (OR, 95% CI: 0.942, 0.914-0.970), inguinal hernia (OR, 95% CI: 0.932, 0.898-0.968), ulcerative colitis (OR, 95% CI: 0.924, 0.860-0.992), and Crohn’s disease (OR, 95% CI: 0.327, 0.211-0.505); neoplasms (n=7): malignant neoplasm of breast (HER-positive) (OR, 95% CI: 1.065, 1.000-1.133), malignant neoplasm of breast (HER2−negative, all cancers excluded) (OR, 95% CI: 1.064, 1.012-1.118), malignant neoplasm of breast (HER2−negative) (OR, 95% CI: 1.061, 1.011-1.114), malignant neoplasm of breast (all cancers excluded) (OR, 95% CI: 1.057, 1.007-1.110), pan cancer (OR, 95% CI: 1.002, 1.000-1.004), ovarian cancer (OR, 95% CI: 1.001, 1.000-1.002), oesophageal cancer (OR, 95% CI: 1.000, 1.000 -1.001); neuropsychiatric system (n=8): Alzheimer’s disease or family history of Alzheimer’s disease (OR, 95% CI: 1.042, 1.005-1.081), father’s history of Alzheimer’s disease (OR, 95% CI: 1.041, 1.005 -1.077), paternal history of Alzheimer’s disease (OR, 95% CI: 1.003, 1.000-1.006), family history of Alzheimer’s disease (OR, 95% CI: 0.923, 0.860-0.990), major depressive disorder (OR, 95% CI: 0.839, 0.730-0.965), hyperkinetic disorders (more controls excluded) (OR, 95% CI: 0.744, 0.615-0.901), mixed disorders of conduct and emotions (OR, 95% CI: 0.731, 0.584-0.916), mixed disorders of conduct and emotions (OR, 95% CI: 0.664, 0.455-0.970); in addition, there were 2 endocrine system disorders and 1 immune disorder: nontoxic diffuse goitre (OR, 95% CI: 1.270, 1.051-1.536), thyrotoxicosis with diffuse goitre (OR, 95% CI: 1.121, 1.017-1.235), and systemic lupus erythematosus (OR, 95% CI: 0.789, 0.643-0.968). SME, WME, and MR-Egger provided reasonably consistent associations (**Supplementary Table S5**).

**Figure 5.**
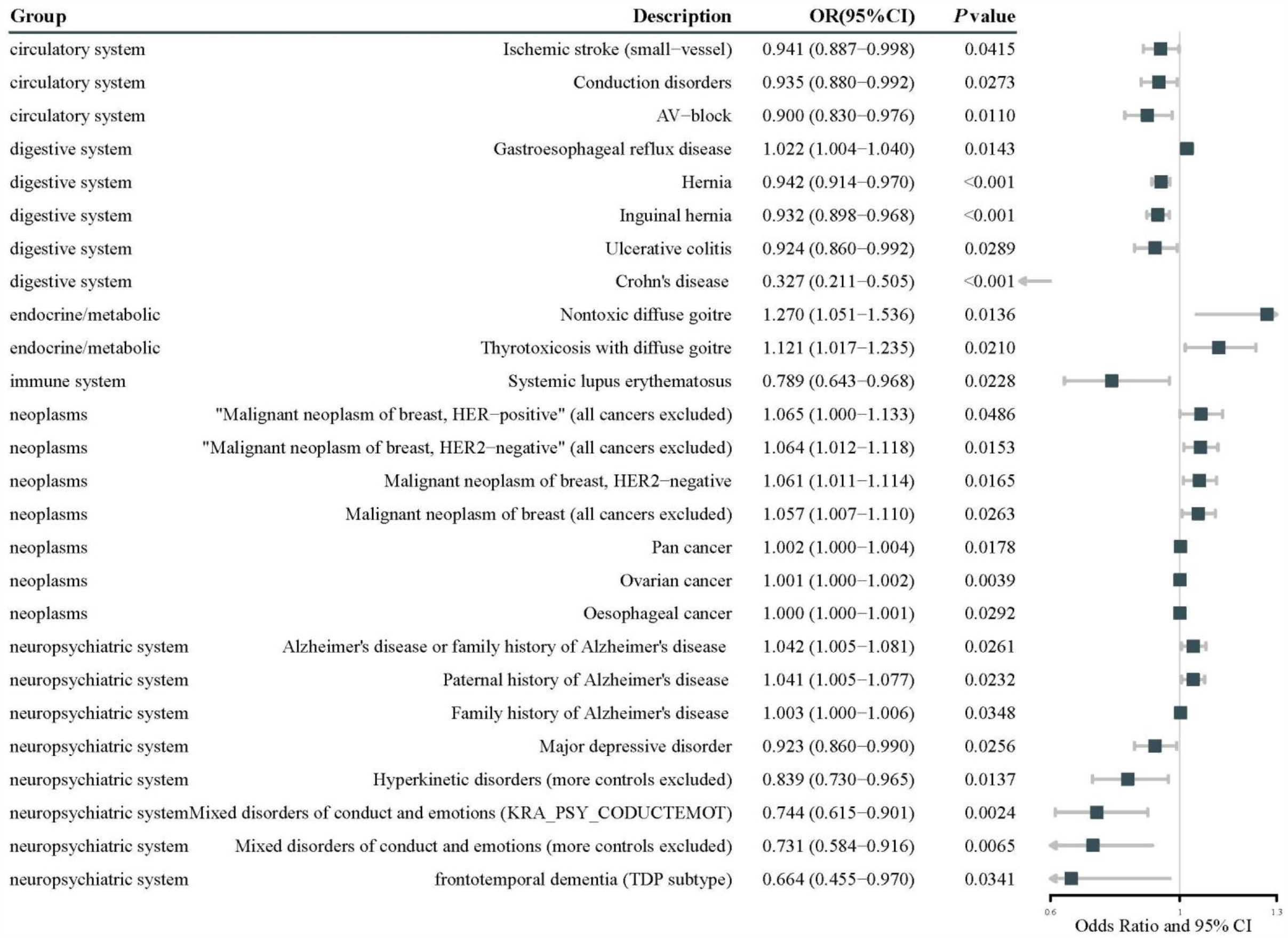
The odds ratios of IVW analysis between severe respiratory COVID-19 and diseases. The forest plot shows a summary of statistically significant (*P*<0.05) results under IVW MR methods; IVW, inverse-variance weighted; OR: odds ratio; CI: Confidence interval;

As shown in **Figure 6**, IVW test found a statistically significant association between hospitalized COVID-19 and 24 diseases, including circulatory system (n=3): ischemic stroke (cardioembolic) [OR 95%CI: 1.092 (1.004−1.186)], ischemic stroke [OR 95%CI: 1.049 (1.001−1.100)], AV−block [OR 95%CI: 0.871 (0.775−0.979)]; digestive system (n=5): gastroesophageal reflux disease [OR 95%CI: 1.035 (1.011−1.061)], hernia [OR 95%CI: 0.930 (0.891−0.971)], inguinal hernia [OR 95%CI: 0.915 (0.869−0.965)], ulcerative colitis [OR 95%CI: 0.694 (0.588−0.819)], Crohn’s disease [OR 95%CI: 0.140 (0.065−0.303)]; neoplasms (n=8): breast cancer [OR 95%CI: 1.139 (1.011−1.283)], malignant neoplasm of breast (HER−positive, all cancers excluded) [OR 95%CI: 1.124 (1.028−1.229)], malignant neoplasm of breast (HER−positive) [OR 95%CI: 1.121 (1.027−1.224)], malignant neoplasm of breast (HER2−negative, all cancers excluded) [OR 95%CI: 1.104 (1.028−1.185)], malignant neoplasm of breast (HER2−negative) [OR 95%CI: 1.100 (1.026−1.179)], malignant neoplasm of breast (all cancers excluded) [OR 95%CI: 1.097 (1.026−1.174)], pan cancer [OR 95%CI: 1.003 (1.000−1.006)], oesophageal cancer [OR 95%CI: 1.001 (1.000−1.001)].

**Figure 6.**
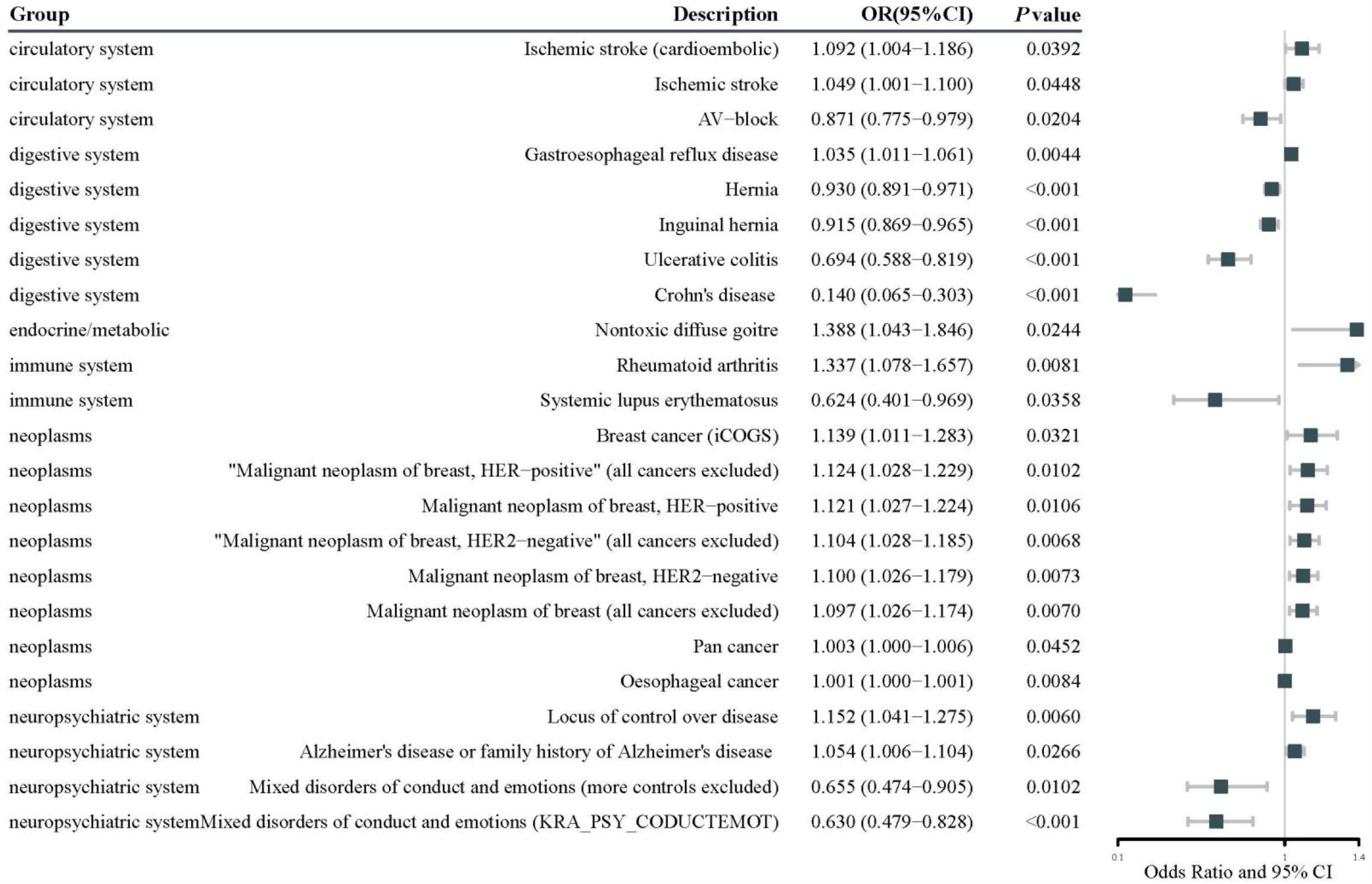
The odds ratios of IVW analysis between hospitalized COVID-19 and diseases. The forest plot shows a summary of statistically significant (*P*<0.05) results under IVW MR methods; IVW, inverse-variance weighted; OR: odds ratio; CI: Confidence interval;

As shown in **Figure 7**, the IVW test found a statistically significant association between susceptibility to COVID-19 and 24 diseases, including circulatory system (n=12): deep vein thrombosis (DVT) of lower extremities [OR 95%CI: 2.392 (1.167−4.902)], DVT of lower extremities and pulmonary embolism [OR 95%CI: 2.052 (1.181−3.564)], venous thromboembolism [OR 95%CI: 1.962 (1.115−3.453)], pulmonary embolism [OR 95%CI: 1.867 (1.161−3.002)], pulmonary heart disease, diseases of pulmonary circulation [OR 95%CI: 1.767 (1.142−2.733)], ischemic stroke (large artery atherosclerosis) [OR 95%CI: 1.405 (1.025−1.927)], coronary heart disease [OR 95%CI: 1.350 (1.006−1.812)], myocardial infarction [OR 95%CI: 1.235 (1.012−1.509)], varicose veins [OR 95%CI: 1.215 (1.001−1.474)], all−cause heart failure [OR 95%CI: 1.140 (1.009−1.287)], conduction disorders [OR 95%CI: 0.780 (0.609−0.997)], AV−block [OR 95%CI: 0.651 (0.467−0.908)]. For full results of the two-sample MR see **Supplementary Table S5**.

**Figure 7.**
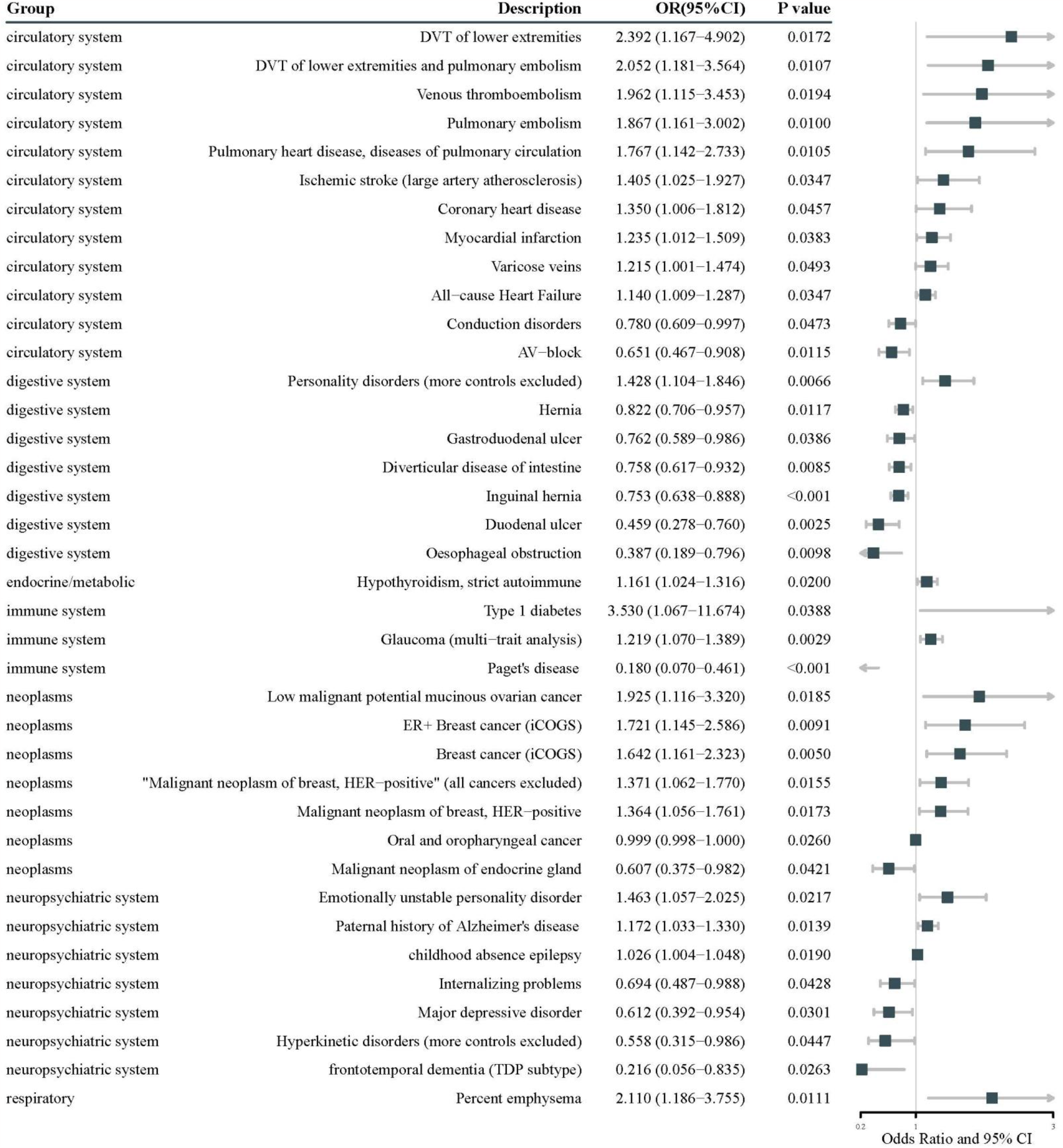
The odds ratios of IVW analysis between susceptibility COVID-19 and diseases. The forest plot shows a summary of statistically significant (*P*<0.05) results under IVW MR methods; IVW, inverse-variance weighted; OR: odds ratio; CI: Confidence interval;

## 4. Discussion

Hypothesis-free PheWAS combined with two-sample MR analysis indicated that genetically determined COVID-19 has a detrimental effect on a broad range of diseases, in particular those impacting the circulatory system, neuropsychiatric system, neoplasms, endocrine/metabolic, and immune system. Results also highlighted the potential impact of COVID-19 on dermatologic, genitourinary, infectious, respiratory, and musculoskeletal disorders. Early detection and management of long COVID-19 could be tremendously beneficial to public health.

COVID-19 as an emerging infectious disease has caused a global pandemic and is spreading at a very rapid rate resulting in an extremely high proportion of people being infected. Clinical symptoms of COVID-19 range from asymptomatic infection to fatal disease [38]. The known potential mechanism is that SAS-CoV-2 enters cells via the angiotensin-converting enzyme 2 (ACE2) receptor [39]. Once inside the body, the virus undergoes replication and maturation, triggering an inflammatory response in which various cytokines are activated and infiltrate a large number of immune cells [40]. ACE2 receptors are present in multiple cell types in the body, including oral and nasal mucosa, lung, heart, gastrointestinal tract, liver, kidney, spleen, brain, and arterial vein endothelial cells, partially explaining the damage caused by SARS-CoV-2 to multiple systems in the body [41]. To date, the generation of long COVID-19 has had numerous effects on the human body [3].

COVID-19-related cardiovascular diseases (CVDs) such as ischemic stroke, coronary heart disease, myocardial infarction, varicose veins, heart failure, and venous thromboembolism occur not only in the early stages of infection but also months later. [42] A study from the UK including 4,182 community patients showed that 13.3% developed at least one persistent symptom 4 weeks after infection, half of which were thought to be of cardiac origin. [16] A recent cohort study by Yan Xie and colleagues based on the national healthcare databases from the US Department of Veterans Affairs found that COVID-19 patients had an increased risk of CVDs compared with controls even among patients who were not hospitalized during the acute phase of infection. [43] In addition, an increased risk of CVDs was observed in the presence or absence of cardiovascular risk factors or preexisting CVDs [43]. These studies suggest that COVID-19 increases the risk of CVDs after acute infection, even in individuals with a low risk of CVDs before COVID-19 infection. Our results suggest that both hospitalized and confirmed neo-coronaries may increase the risk of stroke, and therefore more attention should be paid to patients with neo-coronaries, especially hospitalized neo-coronaries, in terms of stroke prevention. For patients with COVID-19, routine screening for stroke is recommended, including clinical presentation, laboratory tests such as coagulation tests, imaging such as brain imaging, and cerebral angiography. For patients with hospitalized COVID-19, aggressive secondary prevention of stroke should be performed, and anticoagulation therapy is recommended to prevent stroke once laboratory results show a hypercoagulable state.

Significant associations were found between COVID-19 and the neuropsychiatric system using the PheWAS in the UK Biobank. The MR results showed that there was a strong causal effect of COVID-19 on Alzheimer’s disease. Previous studies have suggested that COVID-19 may trigger clinical manifestations of neurodegenerative diseases. [44,45] Recent studies have shown that patients infected with SARS-CoV-2 exhibit reduced brain size, cognitive decline, and damage to brain regions associated with odor. [46] Previous studies have shown that SARS-CoV-2 infection activates TGF-β signaling and oxidative overload and that neuropathological pathways normally associated with AD that lead to tau hyperphosphorylation are activated in COVID-19 patients. [47] However, since most neurodegenerative diseases are late-onset and slowly progressive, current epidemiological studies may not detect such effects to an observable extent. In addition, our MR study found that COVID-19 may increase the risk of neoplasms development. However, no cancer has been reported due to the short observation period of each study. Neoplasms are a serious burden on human health care, thus, it is critical to developing active prevention and treatment solutions for at-risk populations. In addition, the impact of COVID-19 on other systems deserves further evaluation and analysis.

This study has several key strengths. First, a hypothesis-free PheWAS approach was used to gather evidence for a possible causal association between COVID-19 across the range of disease outcomes. Second, the availability of UK Biobank resources enabled the use of information, including hospital episode statistics and mortality data, from over 500,000 individuals. Third, the SNPs used to construct the PRS score were from the most comprehensive COVID-19 GWAS developed to this point.[34] Fourth, the comprehensive analysis strategy included several two-sample univariate MR approaches that could overcome potential pleiotropic effects and avoid overestimating results.

This study also had some limitations. Due to a lack of more publicly available summary-level data, the two-sample MR method could not be used to repeat all results from the PheWAS analysis. In addition, despite the large sample size, UK Biobank is not representative of the general population; [48] however, estimations of risk factor associations are generalizable.[49] Third, the analyses were restricted to participants of White-British descent, thus the generalisability of results to other ethnic groups should be made with caution.

## Conclusions

This study shows a causal effect of genetically determined COVID-19 on a broad range of diseases, in particular those impacting the circulatory system, neuropsychiatric system, neoplasms, immune system, and digestive systems. Early detection and management of long COVID-19 could be a tremendous benefit to public health.

## Data Availability

Details of summary-level data can be found in the supplementary material. Individual-level data that support the findings of this study are available from the UK Biobank (http://www.ukbiobank.ac.uk) (Application ID: 51470).

## Declarations

### Ethics approval and consent to participate

UK Biobank received approval from the UK Biobank Research Ethics Committee (REC; REC reference 11/NW/0382). The ethical approval of summary-level statistics was not applicable but has been approved in the original research.

### Consent for publication

Not applicable.

### Authors’ contributions

FX and KZ have the conception. FX acquire the UK Biobank dataset. KZ did the statistical analyses and drafted the initial manuscript. All authors participated in the interpretation of the results, edited and reviewed the manuscript.

## Acknowledgment

Thanks to UK Biobank and other original research for providing summary-level data resources.

